# Disease-specific plasma protein profiles in patients with fever after traveling to tropical areas

**DOI:** 10.1101/2023.03.10.23287085

**Authors:** Christopher Sundling, Victor Yman, Zaynab Mousavian, Sina Angenendt, Fariba Foroogh, Ellen von Horn, Maximilian Julius Lautenbach, Johan Grunewald, Anna Färnert, Klara Sondén

## Abstract

**Objectives:** Fever is common among individuals seeking healthcare after traveling to tropical regions. Despite the association with potentially severe disease, the etiology is often not determined. Cytokines are soluble mediators dynamically regulated in the response to infection. Measuring cytokines in the blood can therefore be informative to understanding the host-response to infection and can potentially indicate the type of pathogen that causes the disease.

**Method:** In this study, we measured 49 host-response proteins in the plasma of 124 patients with fever after travel to tropical or subtropical regions. The patients had confirmed diagnosis of either malaria, dengue fever, influenza, bacterial respiratory tract infection, or bacterial gastroenteritis, representing the most common disease etiologies. We used multivariate and machine learning methods to assess host-response protein profiles between the different disease groups and healthy control subjects with the aim of identifying disease-associated protein signatures.

**Results:** The host-response varied between disease groups and different combinations of proteins contributed to distinguishing infected patients from healthy controls, and from each other. Malaria displayed the most unique protein signature, indicating a strong immunoregulatory response with high levels of IL10, sTNFRI and II, and sCD25 but low levels of sCD40L. In contrast, bacterial gastroenteritis had high levels of sCD40L, APRIL, and IFN-γ, while dengue was the only infection with elevated IFNα2.

**Conclusions:** These results suggest that characterization of the inflammatory profile of individuals with fever can help to identify disease-specific host responses, which in turn can be used to guide future research on diagnostic strategies and adjuvant treatment.

**Author summary:** Upon infection with a pathogen, the host’s immune system will sense the infection and initiate an immune response. Depending on the type of pathogen and the cells that sense it, the resulting immune response will be different. Fever is a common symptom of infection and it is often difficult to identify the specific pathogen responsible for the disease. In this study, we aimed to characterise and compare circulating inflammation-associated proteins elicited in response to the most common pathogens leading to fever after travel to tropical or subtropical areas. The pathogens included viruses, bacteria, and parasites. Based on the protein signatures, we could observe both disease-general patterns (upregulated in all disease groups) and disease-specific patterns (associated with specific diseases). Malaria displayed the most unique signature and was associated with the upregulation of several immunoregulatory proteins. Possibly in response to a pro-inflammatory response. Dengue fever was also associated with a mix of pro- and anti-inflammatory proteins, while bacterial gastroenteritis had a mainly pro-inflammatory profile. Comparing the protein profiles between diseases indicated unique patterns that could potentially be further developed for clinical use.

## Introduction

Fever is a common symptom of individuals seeking healthcare after returning from travel to tropical areas (1). In a previous study in Sweden, most of these cases were shown to be due to gastroenteritis, malaria, influenza, and dengue virus, but >40% remained with unknown etiology even after healthcare contact (2). Upon further serological analysis of those with unknown etiology, approximately 9% were found to have a likely influenza virus infection, 4% dengue virus infection, and another 4% with Rickettsial infection (2). This highlights the difficulties in travel medicine as the number of pathogens that need to be considered is numerous, and patients with various diagnoses often present with overlapping clinical symptoms (3–6). Moreover, clinical markers such as C-reactive protein (CRP) and white blood cell total and differential counts often provide limited guidance. Pathogens that can give rise to severe disease are important to detect at an early stage, while at the same time, unnecessary antibiotic usage should be avoided (7). Learning more about how immune responses differ between infections with similar clinical presentations could potentially be informative in developing biomarker signatures for disease identification and for a better understanding of host-pathogen interaction and protective immunity.

During infection, a wide range of inflammatory proteins are up- or downregulated in the host’s response to the pathogen. Depending on the infecting pathogen, different types of immune responses are important for efficient control (8). These inflammatory responses tend to be short-lived and self-regulatory as the pathogen is controlled. However, in some instances, the inflammatory response becomes dysregulated leading to severe disease manifestations (9–11). Measuring inflammatory proteins in the blood can therefore help to understand the host inflammatory response to infection and predict disease severity (12–17) or disease etiology (18). Different infections may induce overlapping inflammatory protein profiles and studying a single infection in isolation will make it difficult to understand the pathogen-specificity of the responses and limit the possibility to discern how patterns differ between infections. Recently developed experimental and bioinformatics methods allow for comprehensive analysis of cytokine profiles useful for mapping immunological events as well as for identification of clinically relevant markers of disease and its severity (18–22).

This study provides a comprehensive mapping of inflammatory proteins in the plasma of individuals with a known etiology of infection who presented with fever after travel to tropical or sub-tropical areas. By comparing the inflammatory profile between the different disease groups, we could identify patterns associated with the respective etiologies. These patterns provide insights regarding the pathogen specificity of the host inflammatory response to infection and could potentially be further explored as biomarkers for clinical practice in the future.

## Methods

### Study inclusion

Patients with a history of travel were invited to participate when seeking care at the Emergency Department at Karolinska University Hospital, Stockholm, Sweden. Inclusion criteria were: i) Travel within the past 2 months to a tropical (defined as between latitude 0°-±23.5°) or sub-tropical area (defined as between latitude ±23.5°-±40°), ii) age ≥18 years, and iii) documented body temperature >38°C at the hospital or self-reported fever within the previous 2 days. Blood samples for serum and EDTA plasma isolation were collected from all individuals and aliquots were frozen at –80°C for later analysis. Demographic and clinical data, including microbiological diagnosis after routine clinical investigation, were extracted from medical records and a questionnaire filled in by patients. Five groups of diagnoses were selected for further study: 1) Malaria caused by *Plasmodium falciparum* as defined by microscopy and qPCR, 2) Dengue diagnosed by a positive result in qPCR for dengue virus, 3) Gastroenteritis with fecal culture positive for enteropathogenic bacteria, 4) Influenza diagnosed by positive qPCR for influenza virus in the nasopharyngeal swab, and 5) Bacterial respiratory tract bacterial infections defined as nasopharyngeal culture positive together with chest x-ray with infiltrate or sputum culture positive for a respiratory pathogen together with signs or symptoms indicative of bronchitis or pneumonia. In addition, 13 healthy adult individuals without a current history of travel were sampled as controls. The study was approved by the Swedish Ethical Review Authority (2016/2512-31/2 with amendment 2021-04087). Study participants were provided with written and oral information and written consent was obtained.

In addition to the study inclusion for all tropical fevers, patients with symptomatic malaria had since 2011 been included in a prospective malaria immunology cohort previously investigated for longitudinal parasite persistence (23), cellular aging dynamics (24), parasite-specific antibodies (25,26), B cell responses (27), and immunoregulatory cellular and antibody interplay (19). These individuals were enrolled at the Karolinska University Hospital after confirmed microscopy for *P. falciparum* and providing informed consent. The study was approved by the Stockholm regional ethical committee (2006/893-31/4 with amendments 2018/2354-32 and 2019-03436).

### Multiplex cytokine measurements

Plasma aliquots were thawed at room temperature (RT) and cytokines and chemokines were measured using the LEGENDplex™ pre-defined 13-plex panels according to the manufacturer’s instructions with some modifications. The panels used were; cytokine panel 2, inflammation panel 2, proinflammatory chemokine panel, and the B cell panel (all from Biolegend), measuring 49 unique inflammatory markers in total (**Supplementary Table 1**).

Briefly, 12.5 μl plasma was mixed with 12.5 μl multiplex beads and diluted to 75 μl and incubated for 2h at RT in a plate shaker (700 rpm). The beads were then washed and incubated with 12.5 μl biotin-conjugated marker-specific antibodies diluted to 25 μl with phosphate buffered saline (PBS) for 45 min at RT in a plate shaker (700 rpm). 12.5 μl PE-conjugated streptavidin was then diluted with PBS to 25 μl and added to each well without washing and incubated for 30 min at RT in a plate shaker (700 rpm). The beads were then washed twice, and the fluorescent signal was measured on a BD 4-laser Fortessa using the 488 nm laser (forward scatter vs side scatter) to separate beads based on size and granularity and the 640 nm laser (780/60 filter) to separate beads based on cytokine specificity. The 561 nm laser (582/16) filter was then used to detect the amount of marker signal, translating to the amount of cytokine. The median fluorescent signals were then exported, and the concentration of each marker was determined for each sample based on interpolation from sigmoidal dose-response curves established for each cytokine based on the standards included with the kits. All concentrations below the lower limit of detection (LOD) or above the upper LOD of the assays (as determined by the included standard curve) were assigned a concentration corresponding to the lower and upper LODs, respectively.

### Statistical analysis and machine learning

#### Differential abundance analysis

Data management and statistical analysis were performed using R version 4.3.1. Data visualization was performed using ggplot2 (28). All data on cytokine levels were log-transformed prior to analysis. Spearman’s rank correlation was used to construct correlation networks of cytokine levels within each disease group. For the graphical display of correlation networks, only correlations with absolute values of Spearman’s rho > 0.7 were included. Mann-Whitney-U tests were applied to evaluate differences in the median cytokine levels between the different types of infections or healthy controls. All p-values were adjusted for false discovery rate (FDR) (29). FDR-adjusted *P-*values < 0.05 were considered significant.

#### Discriminant analysis of principal components

Discriminant analysis of principal components (DAPC) was used to examine the multidimensional cytokine data (49 proteins) for specific cytokine profiles/signatures associated with each infection type (30). DAPC is a dimensionality reduction method that identifies the vectors (linear discriminants) in multidimensional space which provides maximum discrimination of groups while minimizing variation within groups. DAPC first transforms the data into uncorrelated variables using principal component analysis and then performs a discriminant analysis on the retained principal components (PCs). Prior to DAPC analysis, all log-transformed values of cytokine levels were scaled and centered by dividing by the median absolute deviation and by subtracting the median, respectively.

#### Feature selection and ROC analysis

The Boruta feature selection algorithm (31) based on binary classification was applied to identify only plasma proteins that contribute significant information for identifying each type of infections from the others and healthy controls. For each infection type, random forest classifiers were then fitted to all the down selected cytokines identified by the Boruta algorithm and the predictive performance was evaluated using receiver operating characteristic (ROC) analysis with 5-fold cross-validation. All data was divided into five parts and in each run, four parts of data were used for model training and the remaining one was used for testing. To further reduce the risk of over-fitting of the classification algorithms, a repeated k-fold cross-validation with 10 repeats and 10 folds was applied for parameter tuning within the model training process. Finally, all ROC plots obtained from five different runs of the model were aggregated into one ROC plot to show the average performance of the model. An imbalanced dataset can result in biased learning algorithms due to differences in the number of individuals in each class. This bias can lead to overly optimistic ROC results that may not be realistic. To address this issue, we also used precision-recall curves to provide a more accurate representation of each model’s performance.

## Results

### Clinical characteristics

In total, 124 patients with self-reported fever seeking healthcare at the Karolinska University Hospital after travel to tropical or subtropical areas were included in the study. A total of 17 patients had dengue fever, 26 had gastroenteritis, 26 had influenza, 49 had malaria, and 6 had bacterial respiratory tract infections. In addition, 14 volunteers were included as healthy controls (**Table 1**). Patients with *P. falciparum* malaria were admitted to the hospital per routine (93.6% of patients) for treatment and observation. Hospital admission was lower in the other groups, with dengue (35.3%), gastroenteritis (38.5%), influenza (19.2%), and respiratory tract infection (66.7%). Among patients with nasal swabs positive for Influenza, 12 patients had Influenza A and 14 had Influenza B by qPCR. Fecal cultures in the group with clinical gastroenteritis revealed growth of either *Salmonella spp.* (n=12) or *Campylobacter spp*. (n=14).

**Table 1.** Clinical characteristics of patients included in the study (n=124)

|  | Dengue<br>n=17 | Bacterial<br>gastroenteritis<br>n=26 | Influenza<br>n=26 | Malaria<br>n=49 | Bacterial<br>respiratory<br>tract<br>infection<br>n=6 | Healthy<br>controls<br>N=14 |
| --- | --- | --- | --- | --- | --- | --- |
| Age, median<br>(IQR) | 37 (33-50) | 36 (28-46) | 37 (31-55) | 38 (30-44) | 57 (39-68) | 30 (26-34) |
| Female sex, n<br>(%) | 6 (35.3) | 12 (46.2) | 15 (57.7) | 9 (18.4) | 4 (66.7) | 10<br>(71.4%) |
| Admitted to<br>hospital, n (%) | 6 (35.3) | 10 (38.5) | 5 (19.2) | 46<br>(93.9) | 4 (66.7) | NA |
| Leukocytes<sup>T</sup>, | 3.1 (2- | 8.8 (4.2-18.9) | 5.5 (3-20) | 5 (1.9- | 8.9 (6.9- | ND |
| ($\times 10^9/L$), median (range) | 7.4) | | | 35) | 11.3) | |
| Platelets<sup>§</sup>, ($\times 10^9/L$), median (range) | 157 (58-346) | 205 (131-549) | 194 (106-442) | 85 (12-210) | 233 (188-367) | ND |
| C-reactive protein<sup>‡</sup>, (mg/L), median (range) | 4 (1-62) | 39 (0-274) | 29 (2-277) | 92 (6-294) | 65 (33-223) | ND |
| Microbiological findings | Dengue virus qPCR positive n=17 | Feces culture positive for Salmonella species: n=12, Campylobacter: n=14 | Influenza qPCR nasal swab positive for Influenza A: n=12, Influenza B: n=14 | Blood smear and qPCR positive for *P. falciparum* n=49. Mean parasitemia 1.7% (range 0.01-17) | Culture positive for *Hemophilus influenzae* Sputum n=4 Nasopharyngeal swab n=2 | NA |

<sup>†</sup>Normal range for leucocyte count $3.8-8.8 \times 10^9/L$
<sup>§</sup>Normal range for platelet count $150-400 \times 10^9/L$
<sup>‡</sup>Normal range for C-reactive protein 0–3 mg/L
N/A – Not applicable
ND – Not done

There was no significant difference in axillary temperature between the infections. They did however differ significantly in leukocyte and platelet counts and levels of C-reactive protein (**Supplementary Figure 1**). Patients with dengue and malaria had significantly lower leukocyte counts compared to bacterial gastroenteritis while dengue also had significantly lower counts compared to influenza and bacterial respiratory tract infection. Platelet counts were significantly lower in the malaria group compared to all other infections. CRP was significantly elevated in all infections except dengue fever which remained close to the reference values. There was no significant difference in CRP values between bacterial gastroenteritis, influenza, and bacterial respiratory tract infection, while malaria was significantly higher than influenza (**Supplementary Figure 1**).

### Different types of infection display unique profiles of inflammatory markers

We measured the levels of 52 inflammatory proteins in the plasma from the study participants and healthy controls using four separate 13-plex suspension bead assays. Two proteins were overlapping between assays, IL12p70 and sCD40L, and had an assay correlation with a Pearson r=0.66 and r=0.82 and both p<0.0001, respectively. One protein (PAI-1) was not quantifiable in all donors and since we could not determine the cause and did not want to introduce potential bias, the PAI-1 data was excluded, leading to a dataset including 49 unique proteins (**Figure 1**). There was a substantial variation in the levels of the proteins among individual donors (**Figure 1A**), however, several disease-specific patterns could be observed (**Figure 1B**). For many of the measured cytokines, levels were highly positively correlated across infections (**Supplementary Figure 2**) as well as within specific infections (**Supplementary Figure 3**).

**Figure 1.**
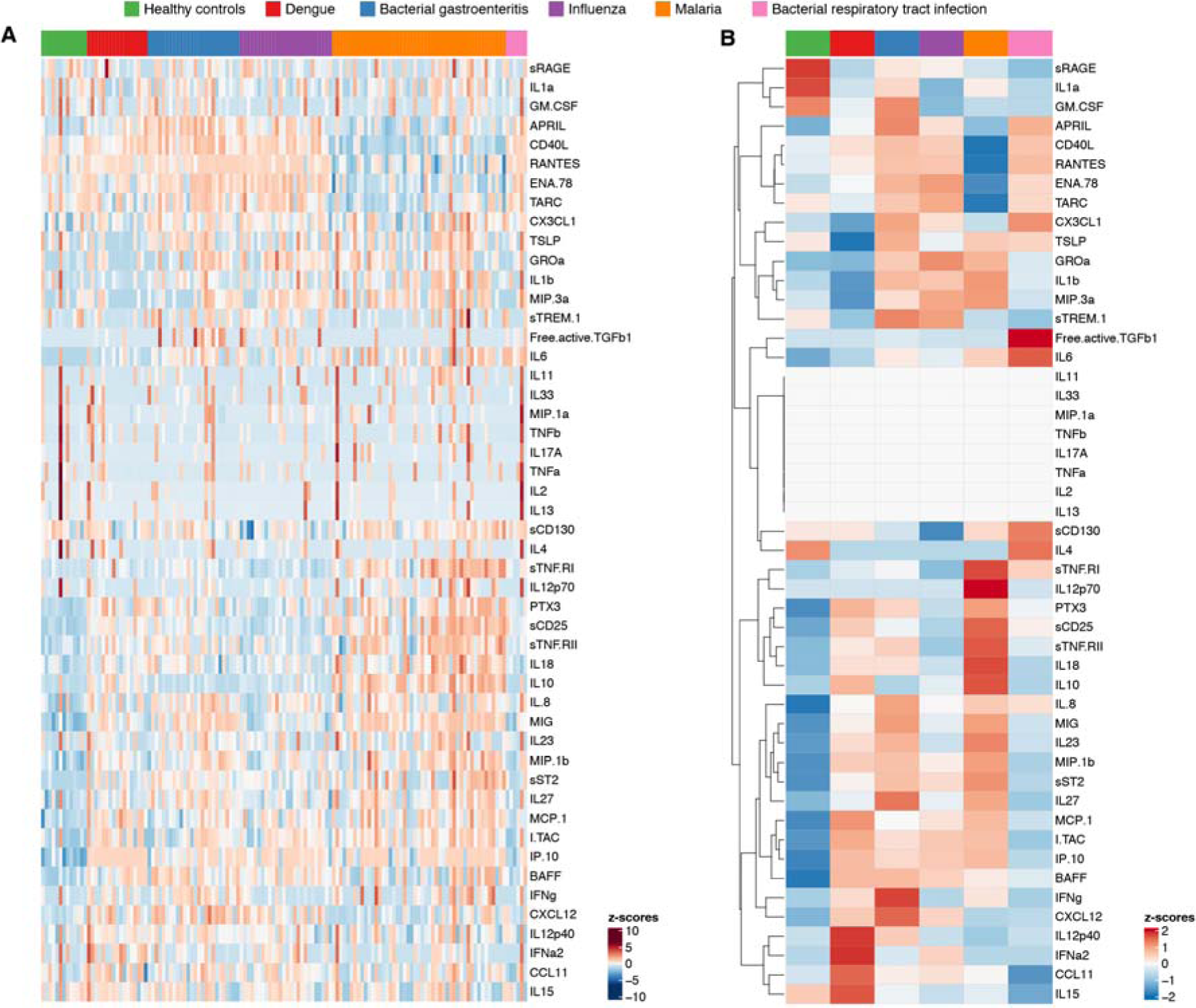
Levels of inflammatory proteins in plasma in different types of infections. (**A**) Heatmap of normalized individual cytokine levels. Each column represents an individual and each row represents a cytokine. Columns are ordered based on disease etiology and rows are ordered using hierarchical clustering based on euclidean distance. (**B**) Heatmap of normalized group-wise median cytokine levels. Each column represents a disease etiology group while each row represents a cytokine.

We next compared the levels of the 49 inflammatory markers between each group and healthy controls using Mann-Whitney-U tests, corrected for multiple testing, to assess if different disease etiologies were associated with up- or downregulation of specific proteins (**Figure 2A)**. Out of the 49 proteins, 29 were significantly up- or downregulated between the different infections or healthy controls (**Figure 2A)**. Compared to healthy controls, dengue patients had a significant increase in 14 out of the 49 measured proteins (logFC>1 and FDR p-value<0.05), while bacterial gastroenteritis led to a significant increase in 18 proteins, and bacterial respiratory tract infection of 3 proteins. Influenza had a significant increase of 12 proteins and the reduction of 1 protein whereas malaria led to significantly increased levels of 17 proteins and reduced levels of 3 proteins. A set of proteins, including BAFF, IL6, IP10 (CXCL10), ITAC (CXCL11), MCP1 (CCL2), MIG (CXCL9), MIP1β, PTX3, sCD25, and sST2 were increased in most infections, potentially indicating quite general markers of febrile illness. There were, however, considerable differences in levels between the groups for some of the markers, such as IP10, where all individuals with dengue had levels above the limit of detection for the assay, and sCD25, which was higher in especially malaria (**Figure 2B**).

**Figure 2.**
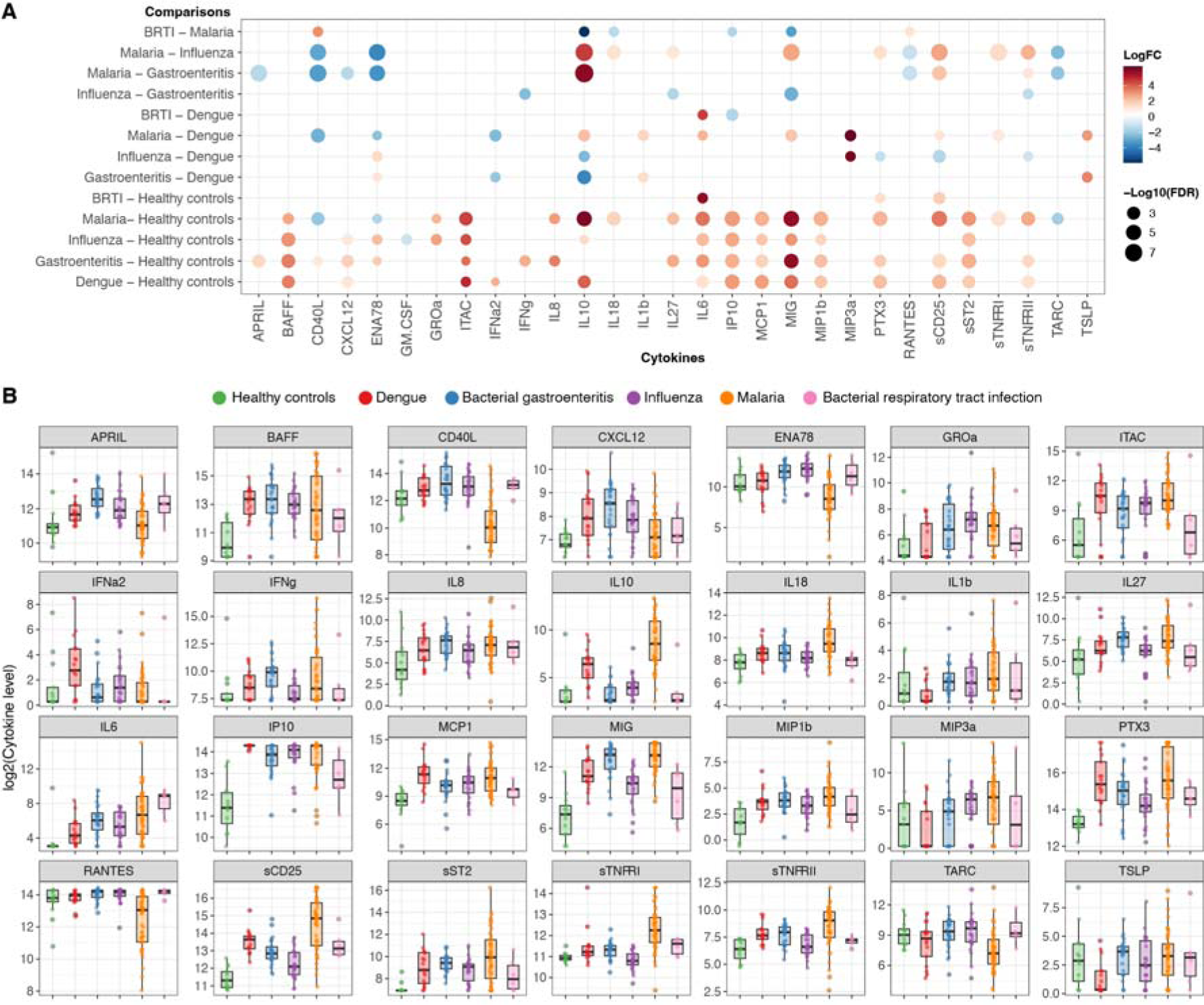
Comparison of inflammatory proteins between disease groups. (**A**) Log-transformed levels (pg/mL) of inflammation-associated proteins were compared between all groups and significant differences, as determined by the FDR-adjusted Mann-Whitney tests, are shown as dots where the size of the dot indicates the p-value and the color indicates a positive (red) or negative (blue) fold-change between the compared groups. BRTI refers to bacterial respiratory tract infection. (**B**) Log-transformed cytokine levels are shown for individual donors in each group. Box plots indicate median and interquartile range.

Some proteins were primarily associated with specific pathogens, such as IFNγ and APRIL which were significantly increased in bacterial gastroenteritis, and IFNα2 which was upregulated in dengue compared with bacterial gastroenteritis, malaria, and healthy controls. IL10 levels were significantly elevated in influenza, dengue, and malaria, with progressively higher levels, while changes to IL18, CD40L, sTNFRI, and TARC were largely specific for malaria. GMCSF levels were slightly lower in influenza compared to healthy controls only (**Figure 2A**). In total, 20 out of the 49 proteins did not differ significantly in any comparison between the different diseases and healthy controls (**Supplementary Figure 4**).

### Immune signatures associated with different infectious diseases

The inflammatory response to infection is complex and is likely affected by the type of pathogen, its virulence, and the anatomical location of the infection. To be able to assess the complexity of the inflammatory response and examine if different diseases were associated with different inflammatory patterns, we used discriminant analysis of principal components (DAPC) (**Figure 3 and Supplementary Figures 5 and 6**). This type of analysis enables us to focus on between-group variability while minimizing within-group variability to identify key markers that segregate the groups included in the analysis (30). The optimal number of PCs to retain for discriminant analysis was determined by cross-validation to be 26. Given the small number of different infections (i.e. clusters) in the data (*k* = 6) all discriminant functions (*k-1*) were retained regardless of their eigenvalue, as they could easily be examined graphically. Data for the first three discriminant functions which explains most of the variation between different infections are presented below in Figure 3, while the full data for all five discriminant functions are presented in Supplementary Figure 5.

**Figure 3.**
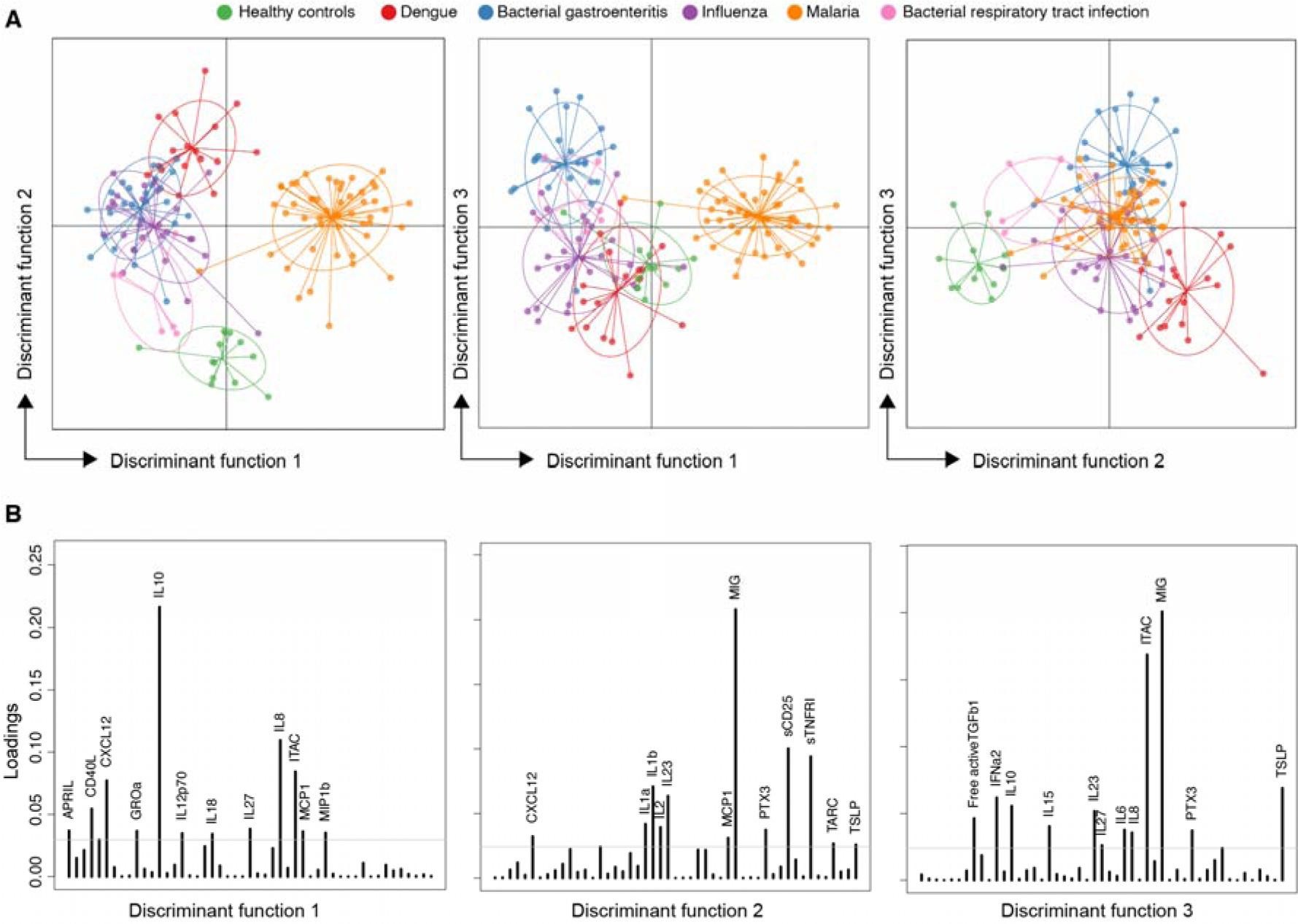
Analysis of marker profiles by discriminant analysis of principal components (DAPC). (**A**) DAPC scatter plots show the separation of groups based on discriminant functions 1-3. (**B**) Proteins associated with separation (loading plots) for each discriminant function. Loading cut-off set to 0.03. Additional data for discriminant functions 4 and 5 are presented in Supplementary Figure 5.

Discriminant function 1 (DF1) provided clear segregation between malaria and the other diseases (**Figure 3A**). Differences in DF1 were largely driven by IL10 levels but also influenced by several other proteins, especially IL6, CXCL12, CD40L, sCD25, and sRAGE (**Figure 3B**). DF2 was instead important for segregating the infected groups from the healthy controls and somewhat from each other. This was largely driven by MIG, but also further influenced by sCD130, IFNα2, CCL11, and IL2. DF3 provided a weaker separation primarily of bacterial gastroenteritis from dengue. This was associated with e.g., IFNα2, as described before, but also TSLP, IL6, MIG, and IL10.

### Selection of proteins indicating different etiologies

To further assess if we could identify an inflammatory signature associated with each type of infection, we used the Boruta feature selection algorithm to identify proteins that contributed significant information for the accurate classification of each disease (**Figure 4**). Since there were only six bacterial respiratory tract infections in the dataset, we did not try to identify a specific signature for this group, although the samples were retained in the dataset when classifying the other groups. For the four remaining diseases, the algorithm identified different numbers of cytokines that provided significant information for classification, indicated by the green color (**Figure 4**). We identified 19, 17, 15, and 21 cytokines for dengue, bacterial gastroenteritis, influenza, and malaria, respectively **(Figure 4).** Several cytokines were selected as important for the classification of more than one pathogen (**Supplementary Figure 7**), such as IL10, MIG, sCD25, and sTNFRI which were selected for all four diseases (**Supplementary Figure 7**).

**Figure 4.**
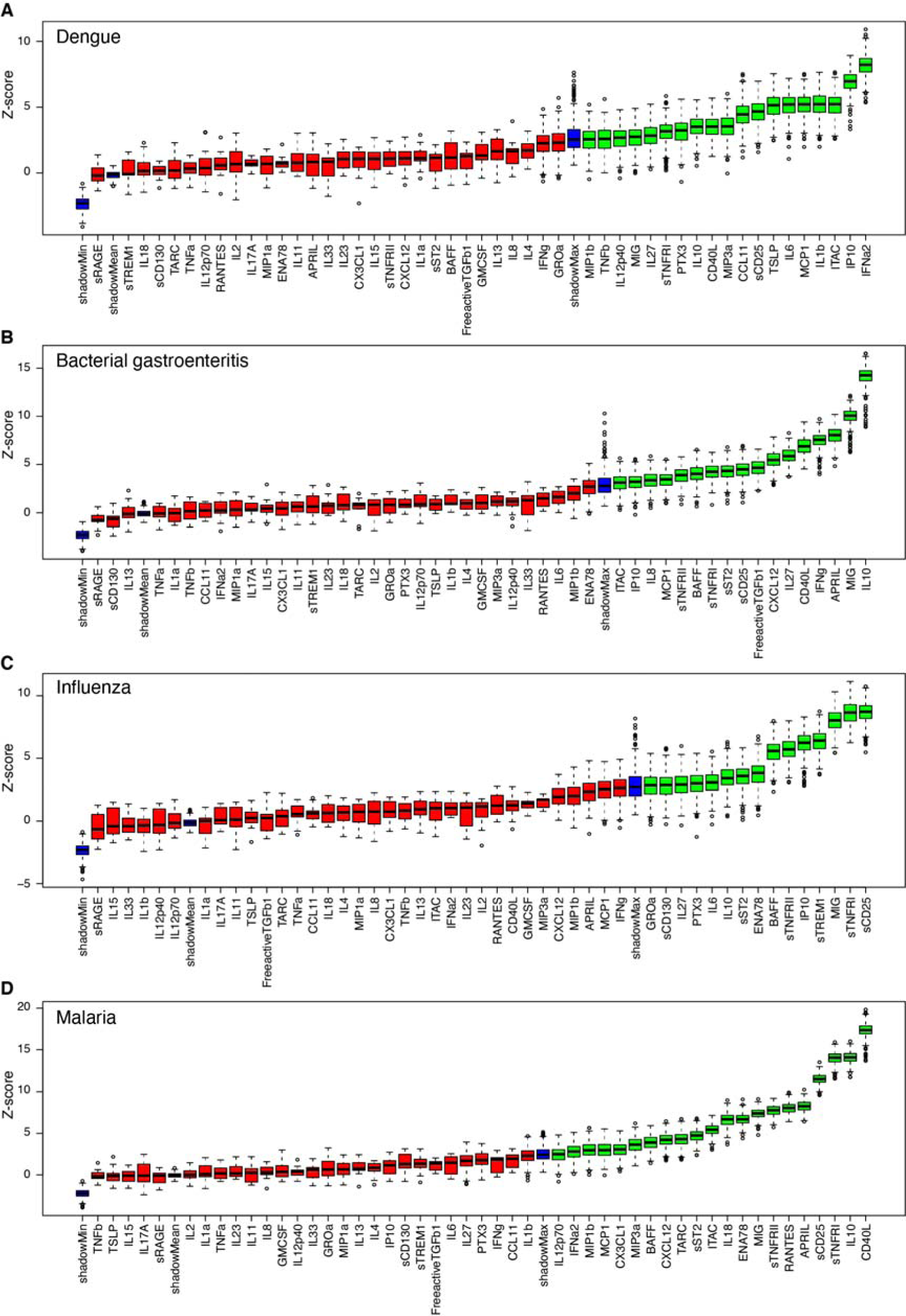
Selecting signatures for detecting individuals infected by each pathogen using the Boruta feature selection algorithm. (**A-D**) Variable importance plots from the Boruta feature selection algorithm fitted jointly to data for all proteins in detecting dengue, bacterial gastroenteritis, influenza, and P. falciparum malaria, respectively. Proteins are ordered from left to right by their importance for classification. The importance measure is defined as the Z-score of the mean decrease in accuracy (normalized permutation importance). Blue boxes correspond to the minimal, average, and maximum Z-scores of shadow features. Red boxes indicate variables not contributing significantly to accurate classification. Green boxes indicate the proteins contributing significantly to the accurate identification of each infection type.

### Performance analysis of the classification of different disease etiologies

Following the Boruta feature selection algorithm, binary random forest classifiers were fitted separately to the data for the proteins selected for each infection (from all individuals) in order to evaluate whether they were informative in identifying individuals with a specific infection type. The best cross-validated classifier performance, as determined by the aggregated classifier area-under-curve (AUC), was observed for malaria, followed by bacterial gastroenteritis, influenza, and then dengue (**Figure 5A**). In malaria, we observed an aggregated cross-validated AUC of 0.97 for a combination of 21 cytokines, while for bacterial gastroenteritis, an aggregated AUC of 0.94 was seen for a set of 15 cytokines. For influenza and dengue, aggregated AUCs of 0.91 and 0.90 were observed from combinations of 17 and 19 proteins, respectively. Overall, all classifiers showed good performance (sensitivity > 0.87 and specificity > 0.72) for detecting each particular infection type using the selected signature.

**Figure 5.**
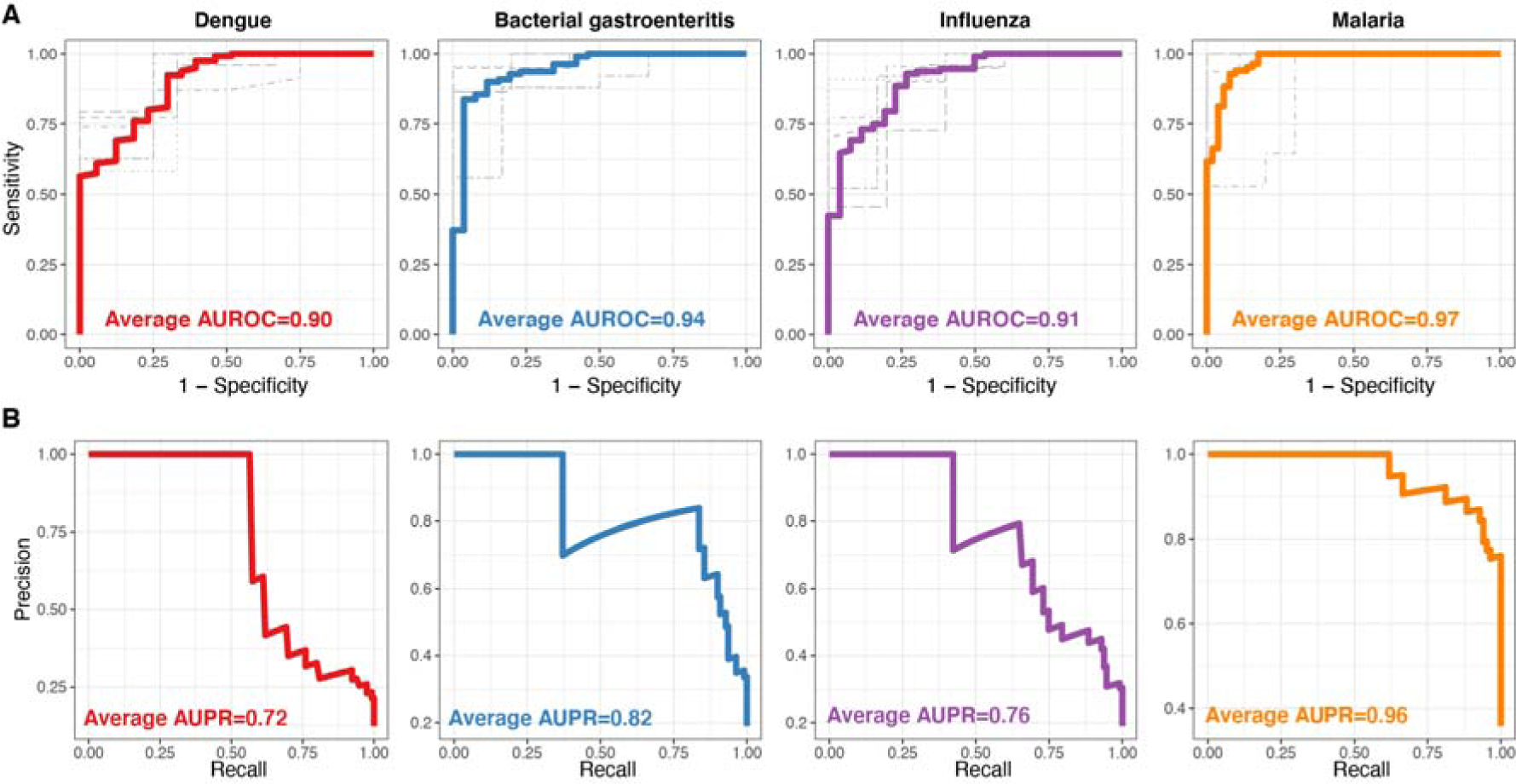
Evaluating performance in identifying individuals infected by each pathogen based on a combination of protein responses. Individual panels display (**A**) cross-validated receiver operating characteristic (ROC) curves and (**B**) aggregated precision-recall (PR) curves for the identification of dengue (red), bacterial gastroenteritis (blue), influenza (purple), and malaria (orange). Random forest classifiers fitted to data on selected proteins that were identified using feature selection for each pathogen. Gray curves in (A) correspond to the ROC curves obtained from the 5-fold cross-validation method and the aggregation of all five ROC curves for the classification of each pathogen is shown with a colored thick ROC curve. The area under the ROC/PR curve (AUC) shows the performance of the classifier. An AUROC/AUPR of 0.5 indicates a classifier that performs no better than random, and an AUROC/AUPR of 1 indicates a perfect classifier.

In our binary classification problem, the goal was to distinguish a specific infection (i.e. the case class) from the other infections and healthy controls (i.e. the control class). In each case, the number of individuals in the control class was greater than the case class, resulting in an imbalanced dataset. To address this issue and report a more accurate model performance, we calculated precision-recall curves (**Figure 5B**). In malaria, the dataset imbalance was very low, resulting in a low bias and a good model performance in terms of both AUC and area under the precision-recall curve (AUPR) (with a sensitivity and precision of 0.9). However, for bacterial gastroenteritis and influenza, with a moderate dataset imbalance, the highest sensitivity and precision were 0.8 and 0.7, respectively. For dengue, we observe the lowest sensitivity and precision, of approximately 0.6, mainly due to a larger class imbalance in the data set (**Figure 5B**).

To further examine to what extent the immune signatures identified above, and the performance of the random forest classifiers, were influenced by differences in inflammatory protein levels between a given infection and healthy controls we repeated the feature selection and classification analysis after first excluding the data for healthy controls. The disease-specific immune signatures as well as the order of importance of different proteins identified in the absence of data for healthy controls differed slightly compared to the original analysis (**Supplementary Figure 8A**). However, the classification performance was largely comparable except for malaria where it was somewhat reduced (**Supplementary Figures 8B**).

## Discussion

Fever is one of the most common symptoms in patients presenting in an emergency setting, especially following travel to tropical- and subtropical regions (32). However, fever is a non-specific symptom associated with many different conditions, some potentially dangerous (6). Despite this, a large proportion of patients seeking care after travel are discharged without identification of the etiological agent causing the disease (2). This could be due to the lack of awareness and/or specific tests for rare pathogens, or difficulty in selecting the accurate test to perform.

Cytokines are immune mediators temporarily produced at high levels during infection, where they provide important functions, such as directly inhibiting pathogen dissemination, stimulating or dampening immune activation, and controlling cellular migration, among other functions (33). The response is generated as a reaction to pathogen-specific patterns and via antigen-specific recognition, potentially making it specific for a given pathogen (34,35). This makes it possible to better understand how the immune system responds to a given infection and potentially predict the type of pathogen based on the inflammatory markers that are increased during the infection. With a broad approach of including viral, parasitic, and bacterial infections in febrile patients we show that many cytokines are up- or downregulated compared to healthy controls and further differ between the infections, making us able to identify disease-specific inflammatory profiles.

We used three complementary approaches to assess the disease-specific inflammatory protein profiles; i) A univariate differential abundance analysis, where protein levels for each individual protein is compared across disease groups; ii) DAPC, an approach that combines both unsupervised and supervised dimensionality reduction techniques, i.e. a principal components analysis (PCA) which reduces dimensionality without considering class labels and a discriminant analysis (DA) that works to maximize the variance between the predefined disease groups, effectively finding the combination of features that best separates these classes, and iii) Random forest classification, a supervised machine learning method, which combines multiple decision trees to predict disease group membership for each given sample, that was used to distinguish each disease from the others. Although the three methods answer fundamentally different questions about the data, they all highlighted a similar set of key proteins that can characterize the main differences in the host inflammatory response towards the different diseases.

Using Boruta feature selection, four proteins (IL10, MIG, sCD25, and sTNFRI) were identified to contribute significantly to stratifying between the four infections included in the analysis, indicating a varied expression in different diseases. These proteins were also highly selected by DAPC for DF1 and DF2. IL10 was among the top three features selected for patients with *P. falciparum* malaria where IL10 was markedly upregulated (also to some extent in dengue virus infection) and for patients with enteric bacterial infection where it remained at baseline levels similar to healthy controls. The increased level of IL10 in patients with malaria is in line with several previous reports (36–38). However, by comparing IL10-levels in malaria with other infections in this study, it becomes clear that the levels reached during acute *P. falciparum* malaria are very high and appear to be a relatively specific hallmark of the disease. In addition to IL10, patients with *P. falciparum* malaria also had especially high levels of sCD25 and sTNFRI. Both these proteins are soluble receptors with sCD25 corresponding to the IL2 receptor and sTNFRI corresponding to the soluble tumor necrosis factor receptor 1 or CD120a, which binds TNF-α. It has been suggested that sCD25 is a marker of T cell activation (39) and has been shown to be increased during different infections, inflammatory diseases, and cancer (40–42). Its purpose remains relatively unclear, but it has been suggested to sequester IL2 and thereby inhibit excessive T cell activation while simultaneously skewing towards the survival of CD25^high^ regulatory T cells (43). sTNFRI is expressed by most cells while sTNFRII is mainly induced in a subset of cells during inflammatory responses. Both receptors are elevated in blood during malaria and correlate with parasitemia in both symptomatic and non-symptomatic infections (44,45) and with the clinical stage and progression of HIV and sepsis (46). sTNFRI is suggested to bind and deactivate excessive TNF to reduce overall inflammation (45). CD40L is another membrane-derived protein belonging to the tumor necrosis factor superfamily. It can have both immunostimulatory and immunoinhibitory effects and soluble CD40L has been associated with the induction of regulatory T cells and immunosuppression in HIV and cancer (47,48). One of the main sources of both membrane-bound and soluble CD40L is platelets (49). In this study, soluble CD40L was somewhat elevated in enteric bacteria compared to healthy controls, but the main difference was a relatively specific and significant reduction in plasma during *P. falciparum* malaria. A potential reason for this strong reduction in soluble CD40L could be thrombocytopenia, as observed in the current study and previous studies of malaria (50,51). Taken together, the high levels of IL10, sCD25, sTNFRI and II, and low levels of soluble CD40L suggest that there is a greatly expanded regulatory or immunosuppressive response generated during acute malaria, perhaps as a counter-effect to the strong immunostimulation coming from high levels of parasites in the blood (52). However, consequently, it has also been proposed that the strong inflammatory and anti-inflammatory response could affect the long-lived adaptive B- and T-cell compartment and reduce the generation of protective immunity (53–55). Although it is difficult to determine the exact effect of this combined response, it is clear that repeated malaria episodes lead to reduced activation of innate immune responses (56–58). This could be an effect of innate training (59,60) or cellular dysregulation (57,61), but could also be influenced by adaptive responses affecting innate activation (19,62,63).

MIG, also called CXCL9, was elevated in all groups compared to healthy controls, potentially working as a general marker of infection. However, the levels were also different between the infections with enteric bacteria and *P. falciparum* malaria having significantly higher levels than both viral and bacterial respiratory tract infections. MIG is induced by IFN-γ and mainly mediates lymphocyte recruitment via binding to its receptor CXCR3 (64). MIG is often also co-expressed with IP10 (also called CXCL10), which was among the top three features selected for dengue virus infection. Like MIG, IP10 was also elevated in all infections, but more so in the dengue group. Since the IFN-γ levels were not higher in dengue compared with enteric bacteria or *P. falciparum* malaria, the increased IP10 levels could come from induction via direct sensing by pattern-recognition receptors (65). In support of this, the level of type I IFN (IFNa2) was elevated in dengue virus infection, but not in the other groups.

A universal marker or combination of markers that could identify specific pathogens would be highly valuable. Clinically available markers, especially the most widely used CRP, WBC, and differential count, can provide some indication of whether acute fever is due to a bacterial or viral infection but they remain relatively unspecific (66–70). In this study, based on 49 markers, we did not observe signatures that were unique to viral or bacterial infections as a group. However, since profiles enriched in several highly important pathogens were identified, these combinations of markers could be further analyzed to improve our understanding of disease-specific immune responses and potentially the identification of disease etiologies.

A strength of this study is that we have explored immune responses in the plasma of individuals with similar symptoms but due to a variety of different microbiological etiologies, contrasting with many studies where only one pathogen and fewer cytokines are studied (71–73). Furthermore, travelers provide a unique opportunity to study host responses following limited exposure and in the absence of re-exposure, in contrast to studies in areas endemic to the disease. The study population was also healthy in general with a median age of 37 years, and therefore with little impact from other chronic diseases or medication, which is often prevalent in patients at the hospital level of care. Conversely, the study also has several limitations. The groups are relatively small for each disease and unbalanced in the number of study participants. They are also not perfectly matched in age or gender. It is therefore important to note that the study primarily has an observational exploratory aim, rather than identifying clinical signatures translating to patient stratification.

In conclusion, our results show that the mapping of plasma protein profiles in febrile patients can identify biomarker combinations that indicate different etiologies. Additionally, we identified cytokines that were uniquely high or low between the infection, indicating different biological functions in the host’s response to infection. Future studies, with larger and more balanced groups with independent training and testing sets, will be important to narrow down the disease signatures to key proteins that could be further developed for clinical tests.

## Acknowledgments

We would like to thank all patients and healthy control individuals who participated in the study. We would also like to express our gratitude to clinicians, nurses and medical students for assisting us with study inclusion. This study was supported by funding from the Magnus Bergvall Foundation (2018–02656), Tore Nilsson Foundation (2018–00608), Swedish Society of Medicine (SLS-934363), Åke Wiberg Foundation (M18-0076), the Swedish Heart-Lung Foundation (20220566) and the Swedish Research Council (2019-01940 and 2021-03706) to CS and grants from Tornspiran foundation to KS and ALF-grants from Region Stockholm to AF. Work with JG was supported by grants from the Swedish Heart-Lung Foundation (20190478), the Swedish Research Council (2019–01034), the Regional Agreement on Medical Training and Clinical Research between Stockholm County Council, and the Karolinska Institutet (20180120).

## Conflict of interest statement

The authors declare that there are no conflicting financial interests.

## Data availability statement

The data will be available upon reasonable request to the corresponding author.

## Code availability statement

All R code for reproducing the analysis are publicly available online at https://github.com/SundlingLab/TF_Cytokines_Study.

## Notes

### Competing Interest Statement

The authors have declared no competing interest.

### Author Declarations

The study was approved by the Swedish Ethical Review Authority (2006/893-31/4 with addendums 2018/2354-32 and 2019-03436). The study was approved by the Swedish Ethical Review Authority (2016/2512-31/2).

### Summary of Updates

First and last author have changed order. Some analysis and text have been added and the discussion has been changed according to suggestions from reviewers. The ms was however rejected and these changes were made to the version now submitted to another journal.

