## Supplemental material for "Disease-specific plasma protein profiles in patients with fever after traveling to tropical areas"

**Running title**: Plasma proteins associated with disease etiology

Supplementary table 1. Proteins measured in the plasma

| **LEGENDplex** | **UniProt** | **Entry Name** | **Gene Names** |
| --- | --- | --- | --- |
| CCL11 | P51671 | CCL11_HUMAN | CCL11 SCYA11 |
| TARC | Q92583 | CCL17_HUMAN | CCL17 SCYA17 TARC |
| MCP1 | P13500 | CCL2_HUMAN | CCL2 MCP1 SCYA2 |
| MIP3a | P78556 | CCL20_HUMAN | CCL20 LARC MIP3A SCYA20 |
| MIP1a | P10147 | CCL3_HUMAN | CCL3 G0S19-1 MIP1A SCYA3 |
| MIP1b | P13236 | CCL4_HUMAN | CCL4 LAG1 MIP1B SCYA4 |
| RANTES | P13501 | CCL5_HUMAN | CCL5, D17S136E, SCYA5 |
| CD40L | P29965 | CD40L_HUMAN | CD40LG CD40L TNFSF5 TRAP |
| GMCSF | P04141 | CSF2_HUMAN | CSF2 GMCSF |
| ENA78 | P42830 | CXCL5_HUMAN | CXCL5 ENA78 SCYB5 |
| MIG | Q07325 | CXCL9_HUMAN | CXCL9 CMK MIG SCYB9 |
| IP10 | P02778 | CXL10_HUMAN | CXCL10, INP10, SCYB10 |
| ITAC | O14625 | CXL11_HUMAN | CXCL11 ITAC SCYB11 SCYB9B |
| GROa | P09341 | GROA_HUMAN | CXCL1 GRO GRO1 GROA MGSA SCYB1 |
| IFNa2 | P01563 | IFNA2_HUMAN | IFNA2 IFNA2A IFNA2B IFNA2C |
| IFNg | P01579 | IFNG_HUMAN | IFNG |
| IL10 | P22301 | IL10_HUMAN | IL10 |
| IL11 | P20809 | IL11_HUMAN | IL11 |
| IL12p40 | P29460 | IL12B_HUMAN | IL12B NKSF2 |
| IL13 | P35225 | IL13_HUMAN | IL13 NC30 |
| IL15 | P40933 | IL15_HUMAN | IL15 |
| IL17A | Q16552 | IL17_HUMAN | IL17A CTLA8 IL17 |
| IL18 | Q14116 | IL18_HUMAN | IL18 IGIF IL1F4 |
| IL1a | P01583 | IL1A_HUMAN | IL1A IL1F1 |
| IL1b | P01584 | IL1B_HUMAN | IL1B IL1F2 |
| IL2 | P60568 | IL2_HUMAN | IL2 |
| IL23 | Q9NPF7 | IL23A_HUMAN | IL23A, SGRF, UNQ2498/PRO5798 |
| IL27 | Q8NEV9 | IL27A_HUMAN | IL27 IL27A IL30 |
| sCD25 | P01589 | IL2RA_HUMAN | IL2RA |
| IL33 | O95760 | IL33_HUMAN | IL33 C9orf26 IL1F11 NFHEV |
| IL4 | P05112 | IL4_HUMAN | IL4 |
| IL6 | P05231 | IL6_HUMAN | IL6 IFNB2 |
| sCD130 | P40189 | IL6RB_HUMAN | IL6ST |
| IL8 | P10145 | IL8_HUMAN | CXCL8 IL8 |
| PTX3 | P26022 | PTX3_HUMAN | PTX3 TNFAIP5 TSG14 |
| sRAGE | Q15109 | RAGE_HUMAN | AGER, RAGE |
| CXCL12 | P48061 | SDF1_HUMAN | CXCL12 SDF1 SDF1A SDF1B |
| sST2 | P30874 | SSR2_HUMAN | SSTR2 |
| FreeactiveTGFb1 | P01137 | TGFB1_HUMAN | TGFB |
| BAFF | Q9Y275 | TN13B_HUMAN | TNFSF13B BAFF BLYS TALL1 TNFSF20 ZTNF4 UNQ401/PRO738 |
| APRIL | O75888 | TNF13_HUMAN | TNFSF13 APRIL TALL2 ZTNF2 UNQ383/PRO715 |
| TNFa | P01375 | TNFA_HUMAN | TNF TNFA TNFSF2 |
| TNFb | P01374 | TNFB_HUMAN | LTA TNFB TNFSF1 |
| sTNFRI | P19438 | TNR1A_HUMAN | TNFRSF1A, TNFAR, TNFR1 |
| sTNFRII | P20333 | TNR1B_HUMAN | TNFRSF1B, TNFBR, TNFR2 |
| sTREM1 | Q9NP99 | TREM1_HUMAN | TREM1 |
| TSLP | Q969D9 | TSLP_HUMAN | TSLP |
| CX3CL1 | P78423 | X3CL1_HUMAN | CX3CL1 FKN NTT SCYD1 A-152E5.2 |

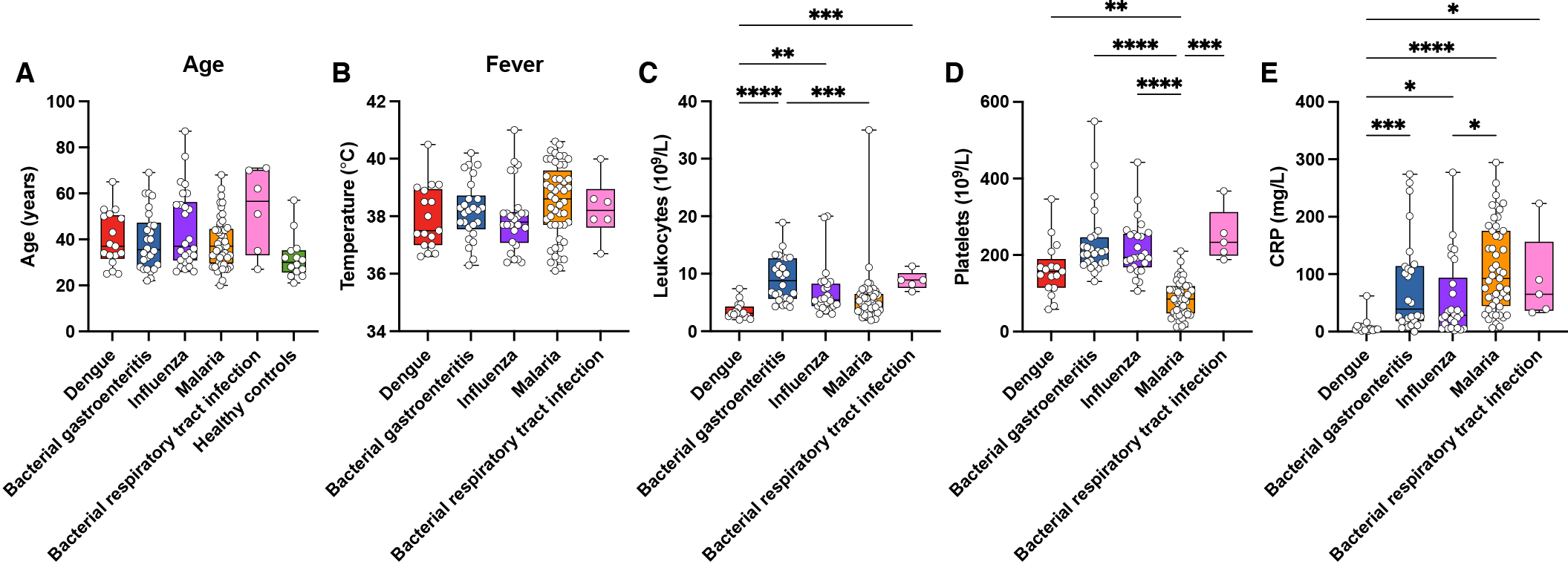

**Supplementary Figure 1.** Clinical characteristics of the study cohort, including (A) Age in years, (B) axillary temperature (°C), (C) leukocyte counts (per 10^9^ per liter blood), (D) platelet counts (per 10^9^ per liter blood), (E) C-reactive protein (mg per liter blood). P-values were evaluated using a Kruskal-Wallis test followed by Dunn’s posttest for multiple comparisons. Only p<0.05 are shown. *p<0.05, **p<0.01, ***p<0.001, ****p<0.0001.

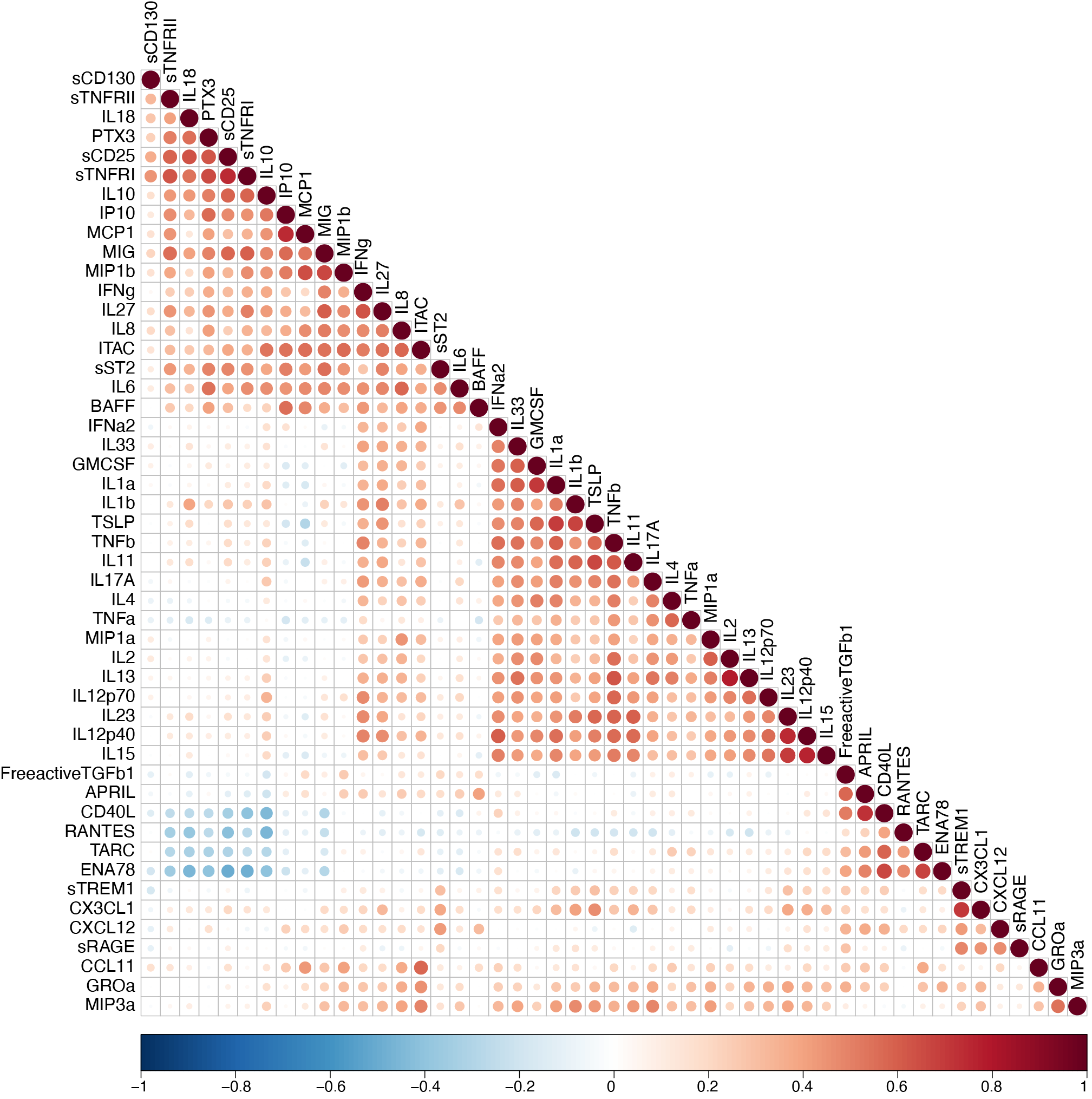

**Supplementary Figure 2.** Correlation matrix heatmap of all pairwise comparisons of cytokine levels. Color intensity reflects the magnitude of the correlation coefficient (Spearmans rho). Dot size corresponds to p-value. Only correlations p<0.05 are shown.

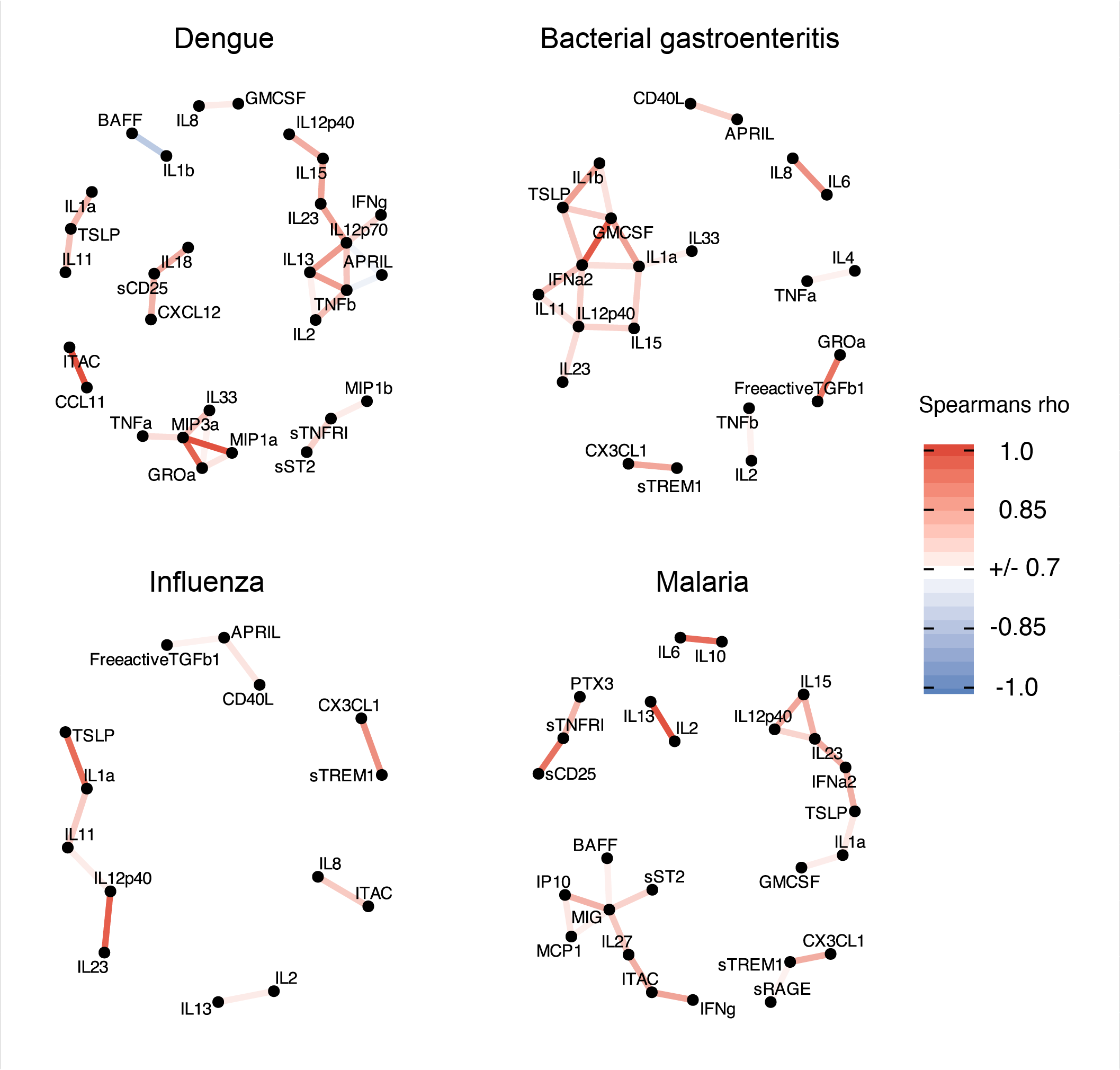

**Supplementary Figure 3.** Correlation network plots of cytokine levels in individuals with dengue (n = 17), bacterial gastroenteritis (n = 26), influenza (n = 26), and malaria (n = 49). Significantly correlating cytokines (rho coefficient of >0.7 or <-0.7 and p<0.05), are displayed as nodes (connected dots). The color intensity of the edges (connecting lines) reflects the magnitude of the correlation coefficient (Spearman's rho). Individuals infected with pulmonary bacteria were omitted from this analysis due to small numbers (n = 6).

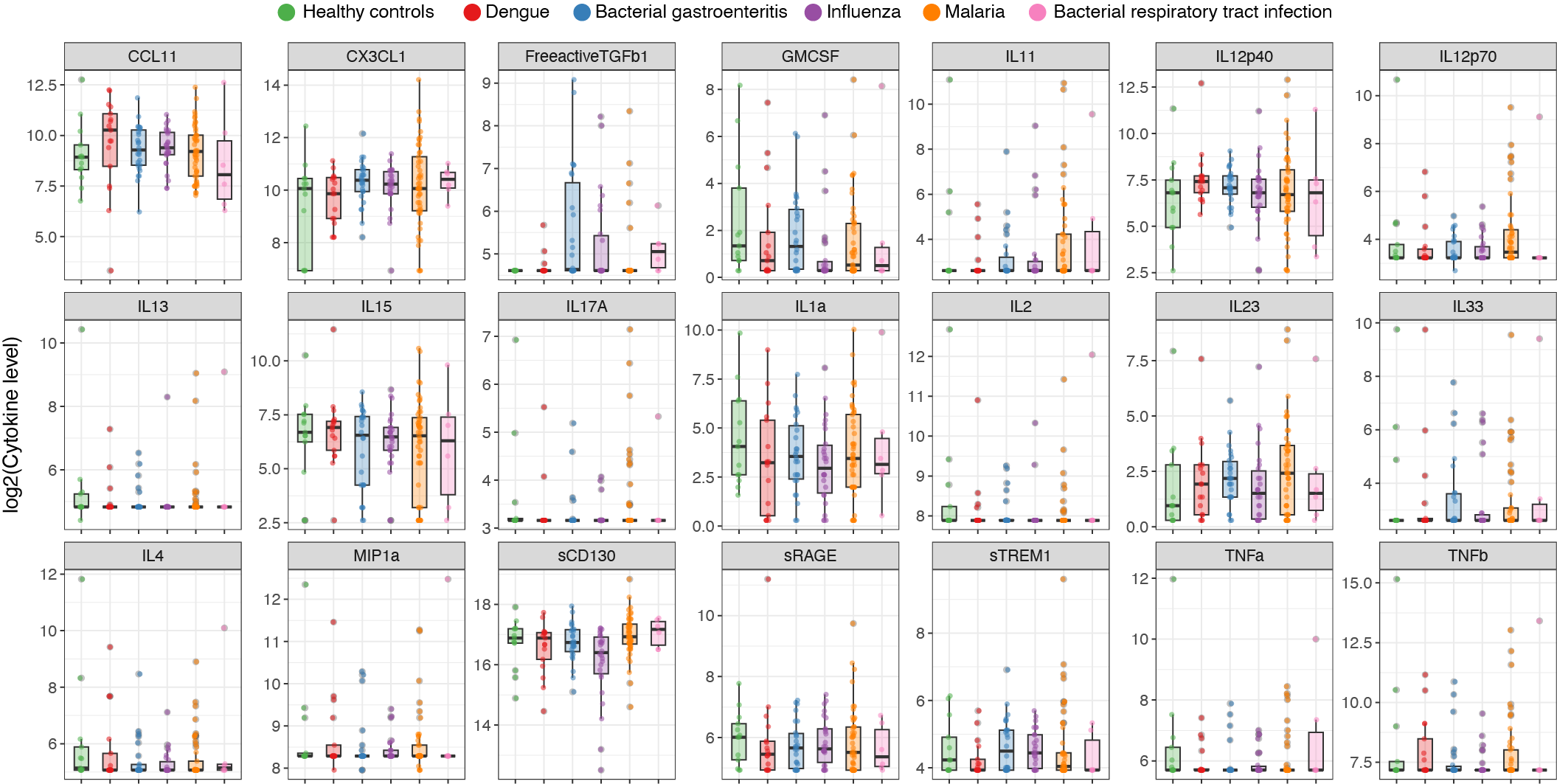

**Supplementary Figure 4.** Levels of proteins with no significant difference between the infected individuals and healthy controls. Box plots show the distribution of log-transformed cytokine levels in different types of infections. Different groups are indicated by color.

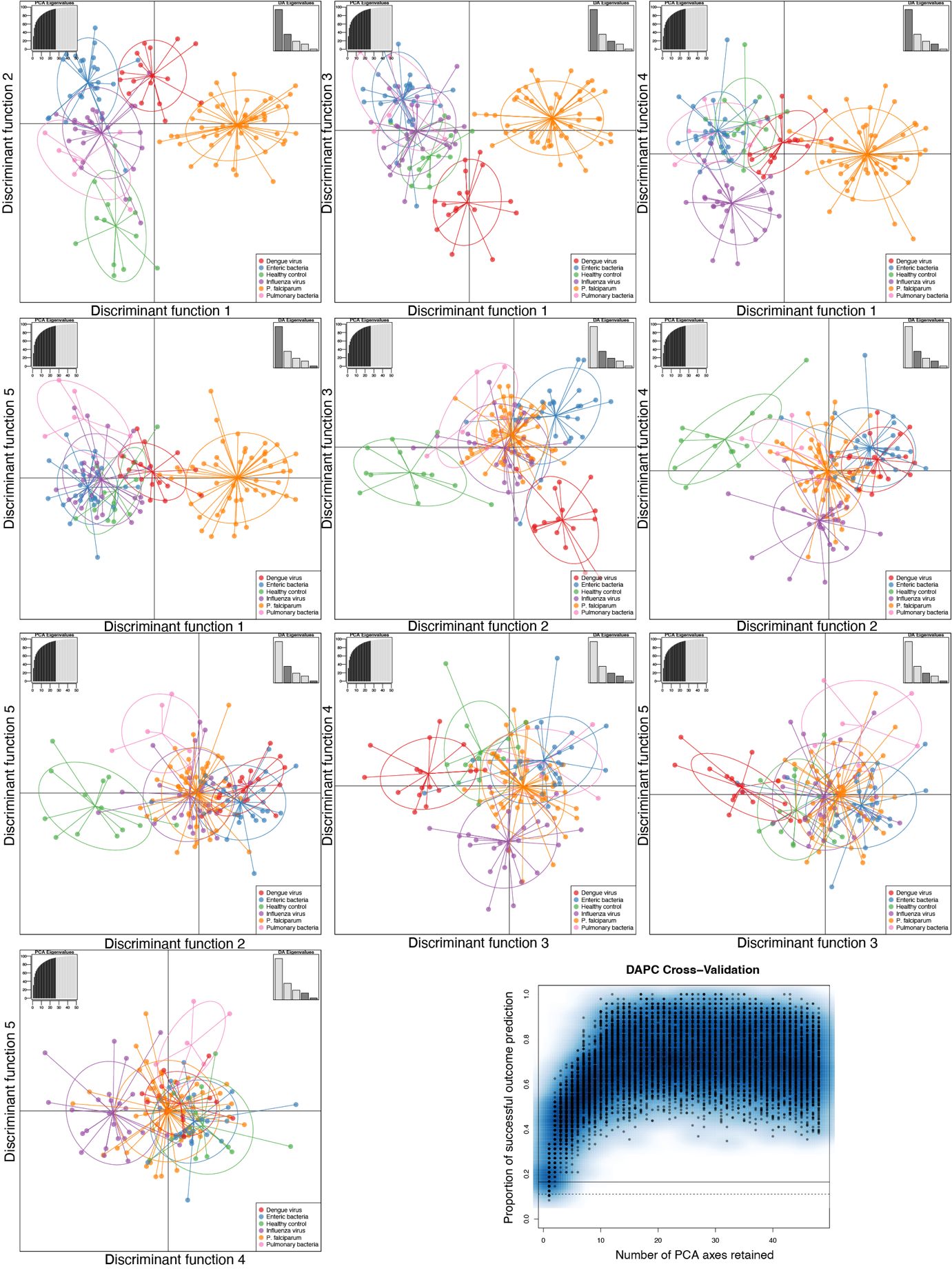

**Supplementary Figure 5**. Discriminant analysis of principal components (DAPC) scatter plots for all pairwise combinations of discriminant functions. The discriminant functions displayed are indicated in the top right corner of each plot panel. For the final DAPC model we retained 26 principal components (PCs) and 5 discriminant functions. The optimal number of PCs to retain was determined using cross-validation as indicated by the lower right panel of the figure.

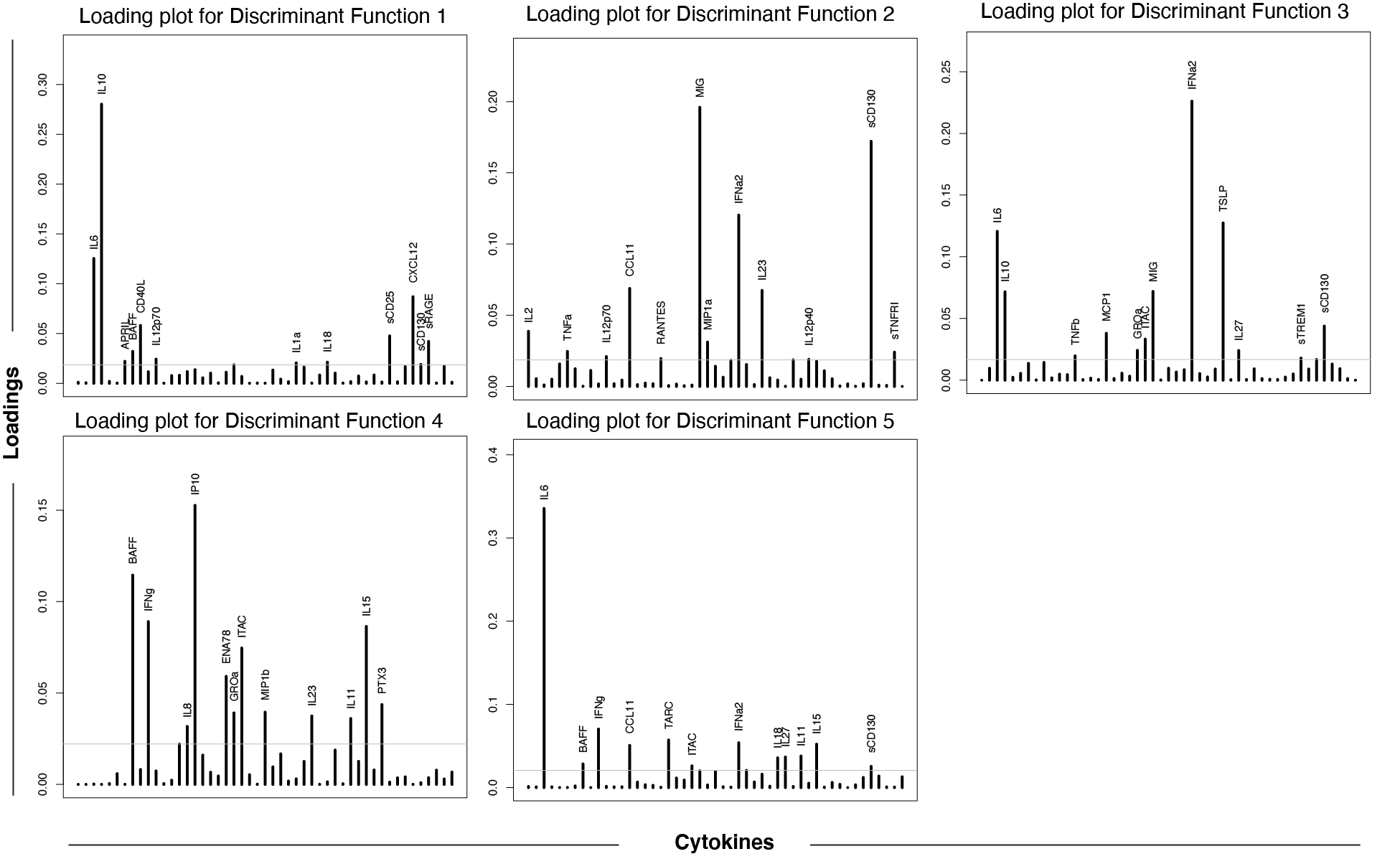

**Supplementary Figure 6.** DAPC Loadings plots. Each panel displays individual variable loadings for a given discriminant function. The bars indicate cytokines with the height of the bar indicating contribution to the discriminant function.

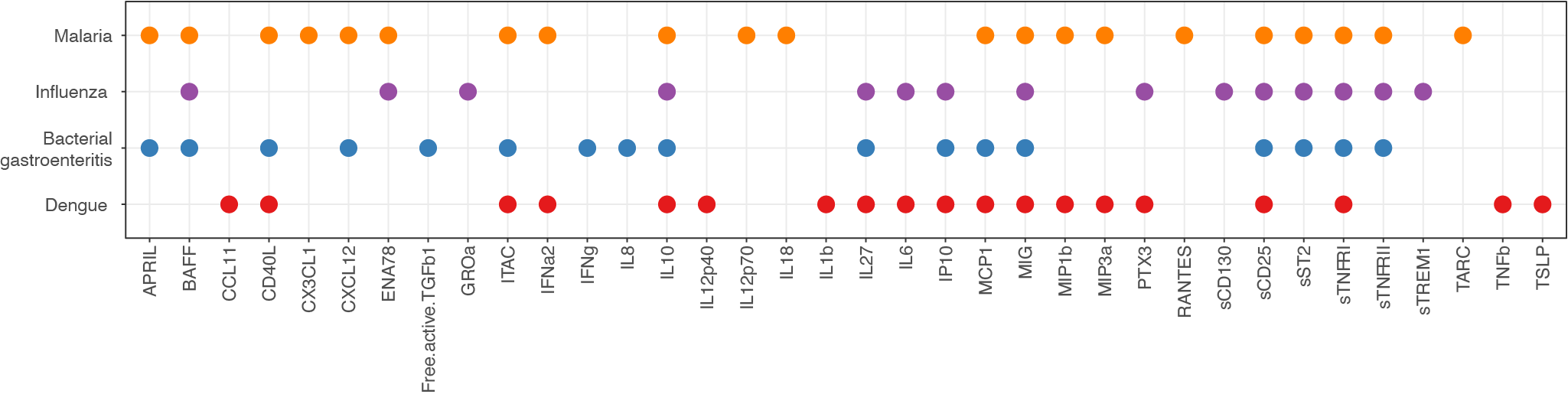

**Supplementary Figure 7.** Overlap of cytokines (x-axis) that are contributing significantly to classification for the different disease groups (y-axis) based on Boruta feature selection.

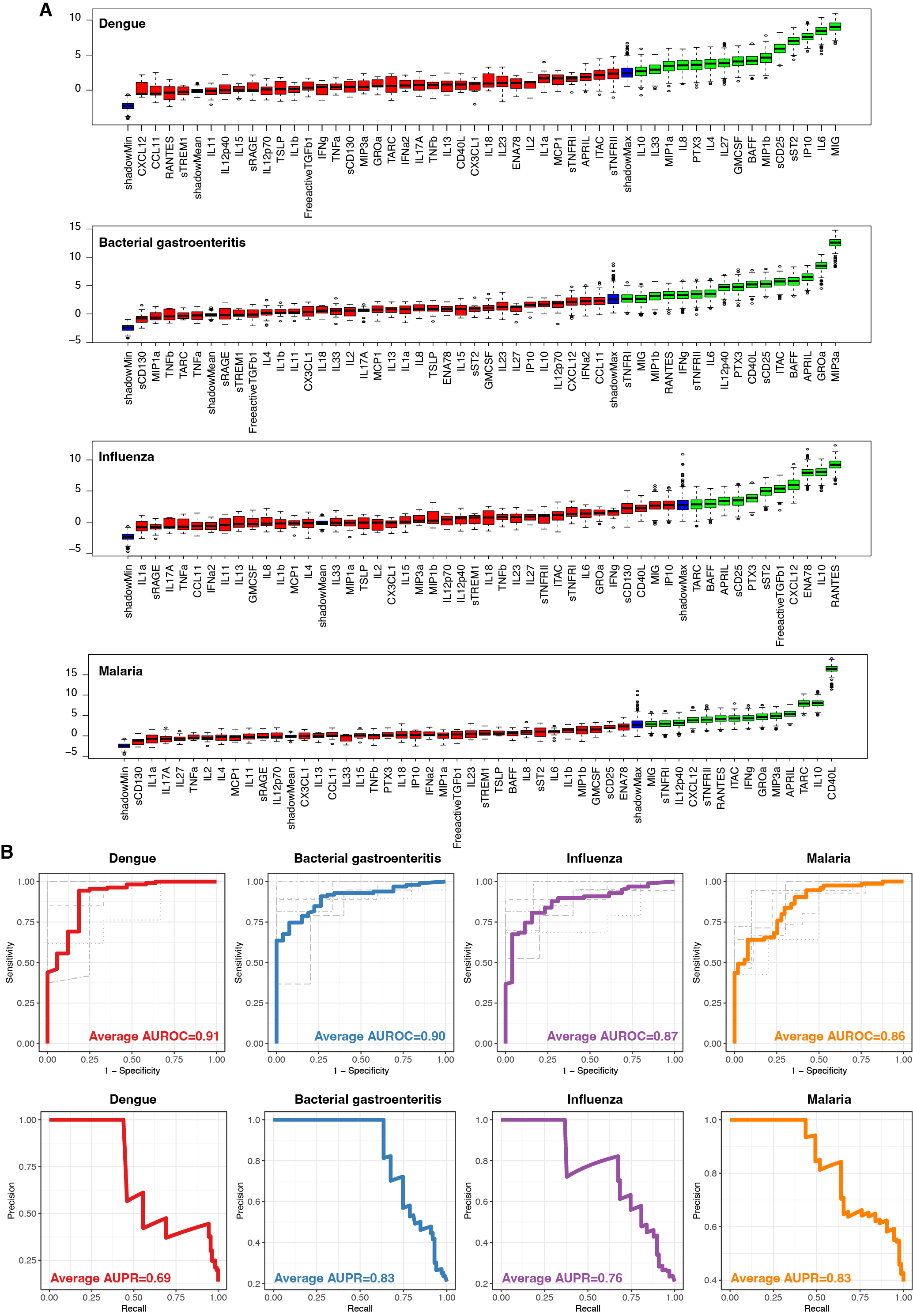

**Supplementary Figure 8.** (**A**) Variable importance plots from the Boruta feature selection algorithm fitted jointly to data for all proteins in detecting dengue, bacterial gastroenteritis, influenza, and P. falciparum malaria, respectively. Data from all infections, but not healthy controls, are included in the analysis (n=124). Proteins are ordered from left to right by their importance for classification. The importance measure is defined as the Z-score of the mean decrease in accuracy (normalized permutation importance). Blue boxes correspond to the minimal, average, and maximum Z-scores of shadow features. Red boxes indicate variables not contributing significantly to accurate classification. Green boxes indicate the proteins contributing significantly to the accurate identification of each infection type. (**B**) Cross-validated receiver operating characteristic (ROC) curves (top panel) and aggregated precision-recall (PR) curves (bottom panel) for the identification of dengue (red), bacterial gastroenteritis (blue), influenza (purple), and malaria (orange). Random forest classifiers from (A) fitted to data on selected proteins that were identified using feature selection for each pathogen. Gray curves in the top panel correspond to the ROC curves obtained from the 5-fold cross-validation method and the aggregation of all five ROC curves for the classification of each pathogen is shown with a colored thick ROC curve. The area under the ROC/PR curve (AUC) shows the performance of the classifier. An AUROC/AUPR of 0.5 indicates a classifier that performs no better than random, and an AUROC/AUPR of 1 indicates a perfect classifier.
